# Placental Pathways: The Impact of Air Pollution (PM_2.5_) Exposure on Pregnancy Outcomes in three Sub-Saharan African Countries

**DOI:** 10.64898/2026.03.10.26348024

**Authors:** Liberty Makacha, P. Tatenda Makanga, Cathryn Tonne, Marie-Laure Volvert, Jovito Nunes, Hawanatu Jah, Esperança Sevene, Moses Mukhanya, Angela Koech, Onesmus Wanje, Anifa Valá, Hiten D. Mistry, Akshdeep Sandhu, Hannah J. Blencowe, Umberto d’Alessandro, Joseph Akuze, Marleen Temmerman, Anna Roca, Jeffrey N. Bone, Yahaya Idris, Laura A. Magee, Ben Barratt, Peter von Dadelszen, PRECISE and PRECISE-DYAD Networks (Supplementary Appendix 1)

## Abstract

**Introduction:** Ambient and indoor fine particle air pollution (PM_2·5_) estimates have been associated with pregnancy complications. We aimed to link direct personal exposure measurements with placenta-mediated pregnancy complications in three sub-Saharan African countries.

**Methods:** We recruited a geographically and energy use stratified sub-sample of 343 rural and urban women who had recently given birth in the PREgnancy Care Integrating translational Science, Everywhere (PRECISE) prospective pregnancy cohort in The Gambia (n = 160), Kenya (n = 105), and Mozambique (n = 78). Individual-level exposure to PM_2·5_ was assessed using high-resolution personal monitoring, during both wet and dry seasons. Minute-level data were summarised as mean and peak daily PM_2·5_ concentrations, and correlated with maternal blood pressure (BP), gestational age at delivery, fetal growth, and stillbirth, in the index pregnancy.

**Results:** 107/343 (32·2%) women experienced pregnancy hypertension, 57/343 (16·0%) women delivered preterm, 203/304 (66·8%) infants with known birthweights were appropriately-grown, and 9/343 (2·7%) infants were stillborn. Higher mean (p=0·012) and peak (p=0·007) exposures were associated with reduced fetal growth velocity, with greater mean exposure associated with small-for-gestational age infants (p=0·016). Greater mean (p=0·017) and peak (p=0·045) PM□.□ exposures were associated with lower birthweight centile. No associations were observed with pregnancy hypertension, pregnancy duration, or stillbirth.

**Discussion:** This study provides exploratory evidence that personal PM_2·5_ exposure is associated with impaired fetal growth in sub-Saharan Africa. Prioritising access to clean fuels, reducing emissions from informal transport and waste systems, and incorporating personal exposure monitoring into maternal health frameworks could yield measurable improvements in birth outcomes and health equity.

## Introduction

A growing global health concern, air pollution is linked increasingly to adverse maternal and perinatal outcomes. Fine particulate matter (PM□.□; ≤2.5 μm), has been associated with hypertensive disorders of pregnancy, small-for-gestational-age (SGA) births, preterm delivery, and stillbirth, often through pathways of placental dysfunction, such as impaired vascular adaptation and oxidative stress.(1-7) These outcomes are interconnected and may reflect shared disruptions to placental development and function.(8)

In Sub-Saharan Africa, PM□.□ exposure has traditionally been attributed to household sources such as biomass cooking, kerosene lighting, and informal waste burning. However, emerging data suggest that substantial exposure also occurs beyond the domestic setting, in outdoor, peri-domestic, and occupational environments.(9, 10) Despite this complexity, epidemiological evidence from sub-Saharan Africa remains sparse, with most studies relying on proxy indicators or ambient estimates that fail to capture the diversity of real-world exposure profiles.

Among the most consistent outcomes linked to air pollution is fetal growth restriction, captured clinically as SGA (birthweight <10^th^ centile for gestational age). SGA is a major risk factor for neonatal morbidity and later-life cardiometabolic disease and may occur with gestational hypertension or stillbirth in settings of placental insufficiency.(11) Personal exposure measurements using wearable sensors offer a more accurate assessment from proxy or modelled exposure estimates. It reflects the cumulative PM□· □ dose individuals experience from all settings, and captures time– activity patterns that influence total exposure burden.(12) Despite its relevance, personal exposure measurement has rarely been implemented at scale in sub-Saharan African pregnancy cohorts,(13) limiting our understanding of how daily pollution realities translate into health risks.

To address this gap, we nested a personal air quality study within the PRECISE (PREgnancy Care Integrating translational Science, Everywhere) pregnancy cohort across The Gambia, Kenya, and Mozambique to examine the relationships between exposure estimates and clinical outcomes related to placental dysfunction, focusing on (i) maternal blood pressure, (ii) gestational age at delivery, (iii) fetal growth velocity, and (iv) stillbirth.

## Materials and methods

In this paper, women are described according to their biological sex assigned at birth.

### The PRECISE Air Quality Cohort

The PRECISE Network recruited unselected pregnant women at the time of booking for antenatal care and followed them until at least six weeks after birth. Women were recruited from community clinics in The Gambia (Farafenni, Iliasa, and Ngayen Sanjal, Farafenni District), Kenya (Mariakani and Rabai, Kilifi County), and Mozambique (Manhiça and Xinavane, Maputo Province). Detailed descriptions of the protocol, database, and cohort profile have been published elsewhere.(14-17) Initially, the plan was to include direct air measurements during index pregnancies; however, due to the impact of the Covid-19 pandemic this was not possible. Therefore, using a stratified random sampling approach (based on key variables such as fuel use behaviours [e.g. cooking, heating, and lighting], and the geographical characteristics of the study areas [e.g. urban vs rural, vegetation index]) to ensure a representative sample across varying personal air pollution profiles, women who had remained where they had lived during their index pregnancy were approached postpartum to participate in the air quality study, that was incorporated into the PRECISE-DYAD (The Gambia and Kenya) and PRECISE-HOME (Mozambique) studies.(17) We planned to recruit approximately 50 women per clinical research site who signed specific informed consent to participate.

### Air Quality Measurement

This protocol has been described elsewhere.(17) In brief, at the initiation of data collection in each of the three study locations, site visits were conducted to provide extensive training to the local field teams. These visits were to support the smooth execution of the data collection process and adherence to the study protocols. During these visits, the field teams were trained regarding the proper management of the wearable sensor devices, including strategies for addressing any technical issues as well as ways to handle any adverse events that might arise. In addition, teams were trained on effective communication with participants, with a focus on ensuring that participants fully understood how to use the sensor bags and follow the study protocols.

Personal exposure to air pollution was assessed using bespoke sensor bags, co-designed by the site and central research teams. Bag design focussed on comfort, discretion, and durability, and ensuring that participants could wear the bags for extended periods without discomfort, while also protecting the sensors from environmental factors such as rain and dust. Before the full deployment of the sensor bags, prototypes were field-tested to ensure they met both the technical and practical requirements of the study. Feedback from participants during early trials highlighted the need for further adjustments in terms of weight distribution and bag size, leading to the final design, which offered improved comfort and functionality.

Air quality research teams constituted study co-ordinators and field co-ordinators from each site, with central coordination from site principal investigators. Sensor bags housed Sensirion SPS030 optical particle counters and other sensors (Dyson Technology Ltd, Malmesbury, UK) that measured PM_2·5_, PM_10_, NO_2_, temperature, humidity, and mobility (via accelerometery and geolocation) in real-time. The sensors were programmed to collect data at 1-minute intervals, providing highly granular insights into individual exposure patterns throughout the day. Data collection was conducted from the same participants during both the dry and wet seasons, with up to 10 participants recruited per week, over a six-week period in each season (‘campaign’). While the exact number of participants varied weekly, this recruitment strategy ensured that the data collected spanned the breadth of each season, capturing the seasonal variations in air pollution exposure across the study sites.

Participants were instructed to wear the sensor bags continuously throughout the day, from the moment they began their activities until they went to bed. During the night, participants were advised to place the sensor bag within one metre of their usual sleeping area to ensure continued capture of overnight exposure. This approach ensured comprehensive, 24-hour exposure data, reflecting both active and passive environmental exposures over the course of the study period.

For purposes of safeguarding data integrity and confidentiality, all data collected by the sensors were encrypted at the point of collection and uploaded to a secure server at Imperial College London after each cycle of data collection at each site. Data remained encrypted throughout the storage process and were only decrypted at the point of analysis, adhering to strict confidentiality protocols to protect participant information.

### Outcomes

The outcomes of interest were related to four key outcomes: (i) blood pressure; (ii) gestational age at delivery; (iii) attained size-at-birth (as a proxy for fetal growth velocity); and (iv) stillbirth.

For blood pressure, we assessed the relationship between PM_2·5_ exposures and gestational hypertension and pre-eclampsia, both defined according to the International Society for the Study of Hypertension in Pregnancy.(18) In brief, gestational hypertension was defined as hypertension (systolic blood pressure [sBP] of at least 140 mmHg or diastolic blood pressure [dBP] of at least 90 mmHg) first diagnosed at or after 20 weeks 0 days gestation, without features of pre-eclampsia. Pre-eclampsia was defined as gestational hypertension with either significant proteinuria, evidence of end-organ complications (e.g. kidney or liver), or evidence of placental dysfunction (e.g. fetal growth restriction); it can be superimposed on chronic hypertension. In addition, we assessed the relationship between PM_2·5_ exposures and mean arterial pressure, defined as dBP + 1/3 of the pulse pressure (sBP– dBP).

For gestational age at delivery, we assessed the relationship between PM_2·5_ exposures and term (birth at or after 37 weeks 0 days), late preterm (birth at 34 weeks 0 days to 36 week 6 days), moderate preterm birth (birth at 32 weeks 0 days to 33 weeks 6 days), and very preterm birth (birth before 32 weeks 0 days). In addition, we assessed the relationship between PM_2·5_ exposures and gestational age at delivery.

We assessed the relationship between PM_2·5_ exposures and very large size for gestational age (birthweight greater than the 97^th^ centile for gestational age) large for gestational age (birthweight at the 90·1^th^ to 97^th^ centile for gestational age), appropriate for gestational age (birthweight at the 10^th^ to 90^th^ centile for gestational age), small for gestational age (birthweight at the 3^rd^ to 9·9^th^ centile for gestational age), and very small for gestational age (birthweight less than the 3^rd^ centile for gestational age) as proxies for fetal growth. In addition, we assessed the relationship between PM_2·5_ exposures and birthweight centile. Sex-specific birthweight centiles were defined according to the INTEGROWTH-21^st^ standards.(11)

For stillbirth, we assessed the relationship between PM_2·5_ exposures and stillbirth, defined as the birth of an infant of at least 20 weeks 0 days gestation or 500 g birthweight without vital signs.(19)

### Statistical Analysis

Our PM_2·5_ exposure metrics of interest were mean and peak exposures, computed per participant per season using 1-minute resolution data collected over five consecutive monitoring days.PM□.□. Peak exposure was defined as the 75th percentile of minute-level PM□.□ values per participant per season. For descriptive analyses, we used Mann-Whitney U tests, Kruskal-Wallis non-parametric ANOVA tests, and Dunn’s post-tests for grouped continuous variables. To examine the relationships between PM_2·5_ exposures and continuous variables (e.g. mean arterial pressure), we examined both linear regression and generalised additive model (GAM) first-order splines. All analyses were performed in R statistical software version 4·4·3 (R Foundation for Statistical Computing, Vienna). Non-overlapping interquartile ranges and p < 0·01 were used for statistical significance, to correct for multiple comparisons. For regression analyses, results were considered significant if both linear regression and GAM p-values were < 0·05.

### Ethics Approvals

Approval for the PRECISE-DYAD study was obtained from King’s College London (Ref HR-20/21-19714), The Gambia Government/MRC Joint Ethics Committee (Ref 22843), Aga Khan University, Nairobi Institutional Ethics Review Committee (Ref 2021/IERC-08) and University of British Columbia (Ref H20-02769). Approval for the PRECISE-HOME study was obtained in King’s College London (Ref HR-17/18–7855), the Mozambique Ministry of Health, National Bioethics Committee for Health (CIBS-CISM/105/2021). Approval for the PRECISE study was obtained in King’s College London (Ref HR-17/18–7855), Aga Khan University Hospital (Ref 2018/REC-74), The Gambia Government/The Medical Research Council, The Gambia Joint Committee (Ref SCC 1619), and the Mozambique Ministry of Health, National Bioethics Committee for Health (545/CNBS/18).

## Results

The PRECISE Air Quality Cohort comprised 343 pregnant participants recruited from The Gambia, Kenya, and Mozambique. Exposure monitoring was conducted over 328 unique days between March 2022 and January 2023, spanning both dry and wet seasons, and enabling the capture of a wide range of settings, environmental conditions, and pollution profiles (Table 1). Generally, women were in their twenties, and of variable parity, educational status, risk of poverty, and rurality, with higher-than-anticipated burdens of pregnancy hypertension (107/343 [32·2%]), preterm birth (57/343 [16·6%]), small-for-gestational infants (74/343 [24·3%]; 39 [11·4%] birthweights were missing due to reduced research activities during the Covid-19 pandemic), and stillbirth (9 [2·6%]).

**Table 1.**
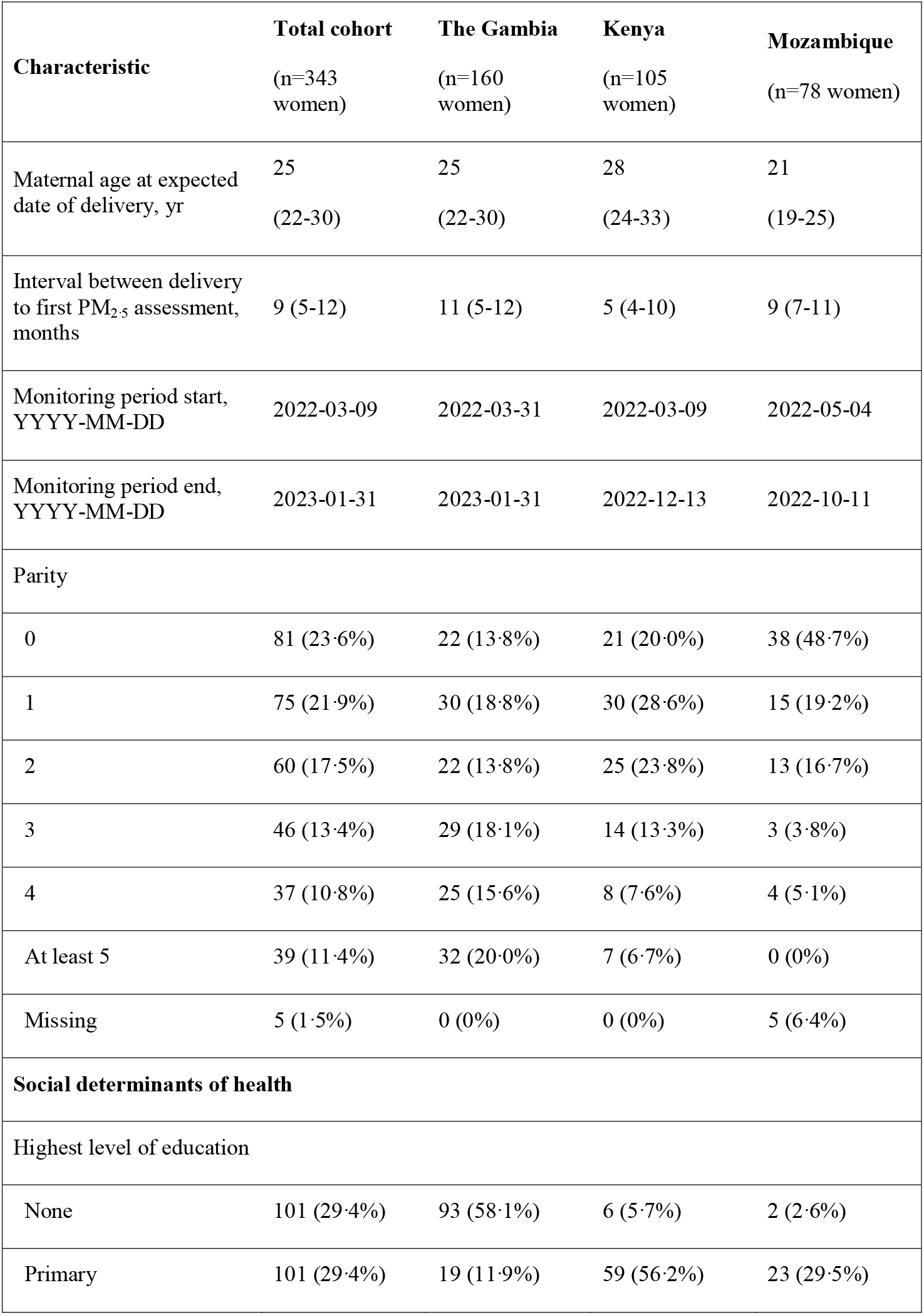
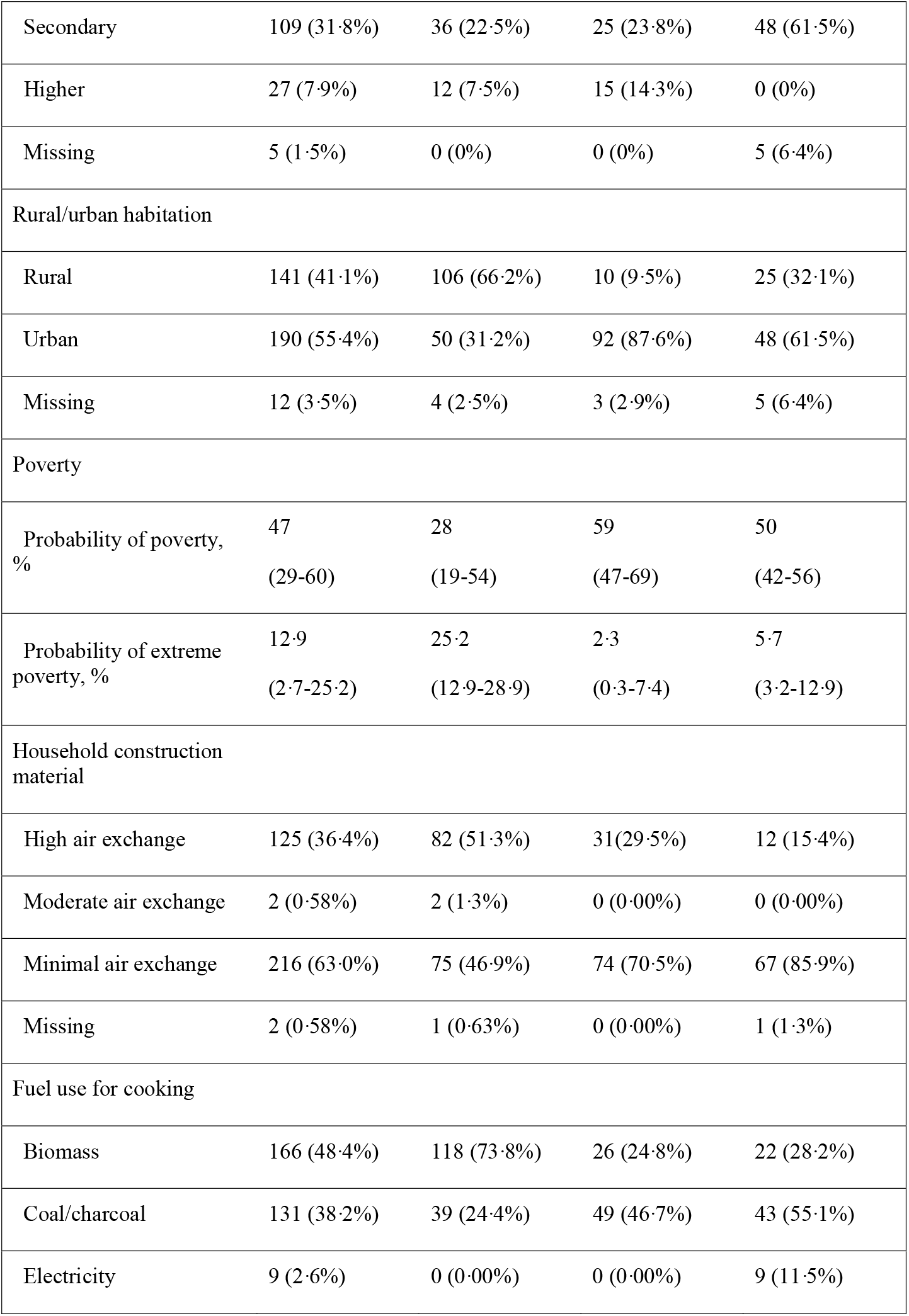
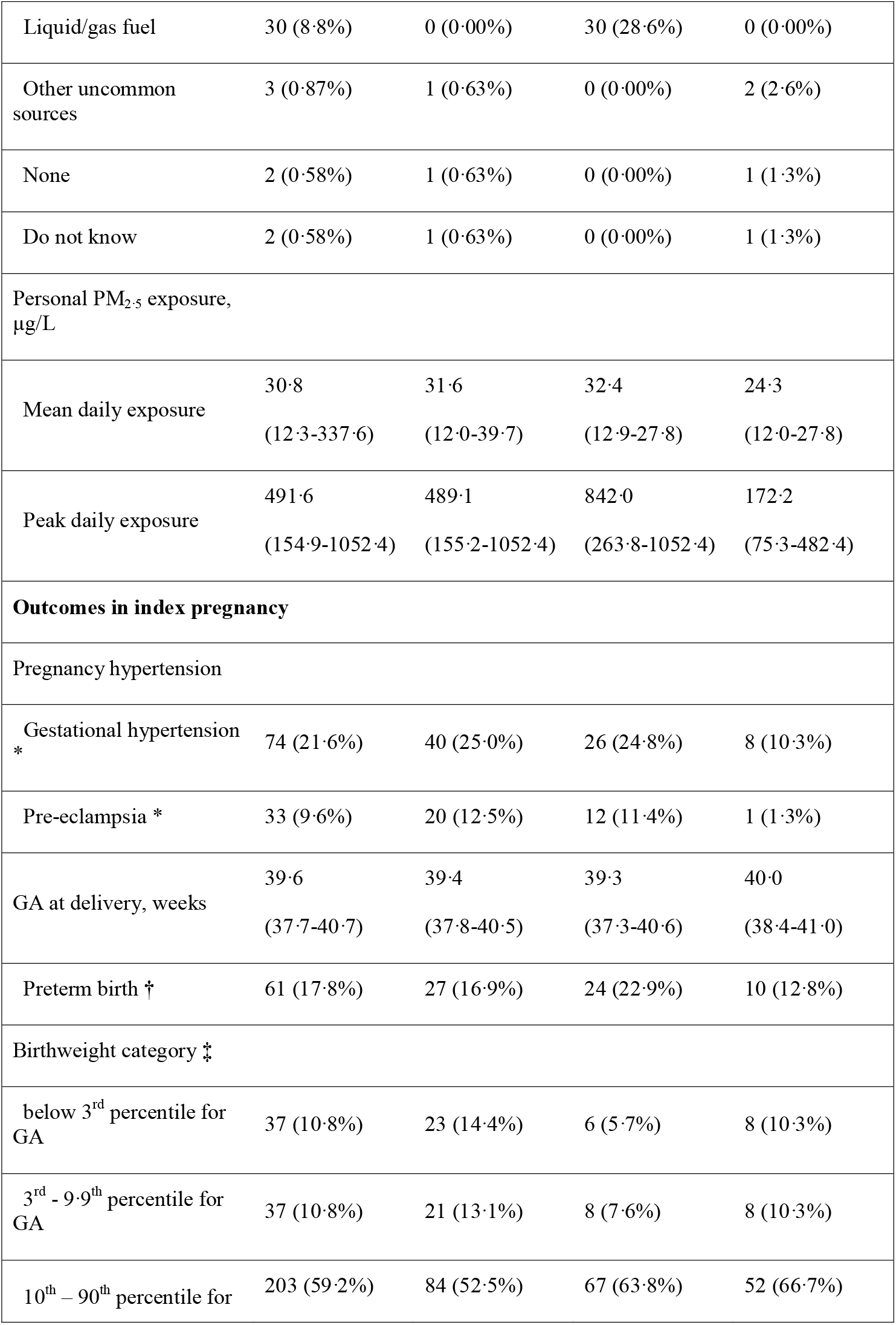
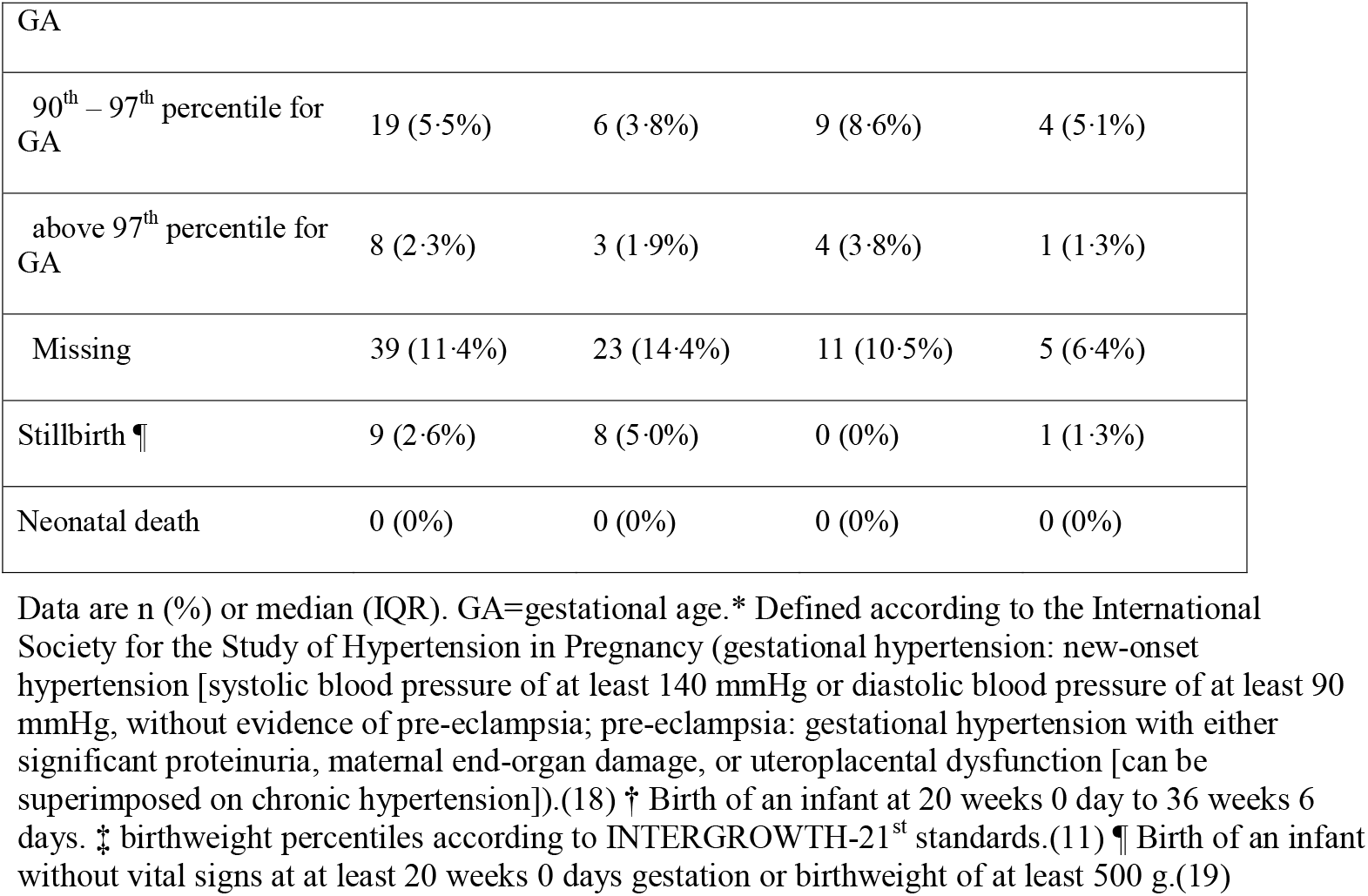
Characteristics of the PRECISE air quality cohort.

Personal exposure to fine particulate matter (PM_2·5_) was greater than the upper limit of World Health Organization (WHO) air quality guidance (i.e. 5 µg/m^3^ for annual exposures; 15 µg/m^3^ for short-to-long term exposures) across all sites,(20) but highly variable between participants. Mean PM_2·5_ concentrations ranged from 24·3 µg/m^3^ in Mozambique to 32·4 µg/m^3^ in Kenya. Peak PM_2·5_ concentrations ranged from 172 µg/m^3^ in Mozambique to 842 µg/m^3^ in Kenya.

In terms of maternal blood pressure (figure 1), for mean PM_2·5_ (Kruskal-Wallis p = 0·017) and peak (Kruskal-Wallis p = 0·030) exposure there were trends towards variation between groups. Increasing peak PM_2·5_ concentrations were associated with higher mean arterial pressure in wet seasons; linear regression p = 0·022, GAM p = 0·0067 (supplementary figures 1 a-c).

**Fig. 1.**
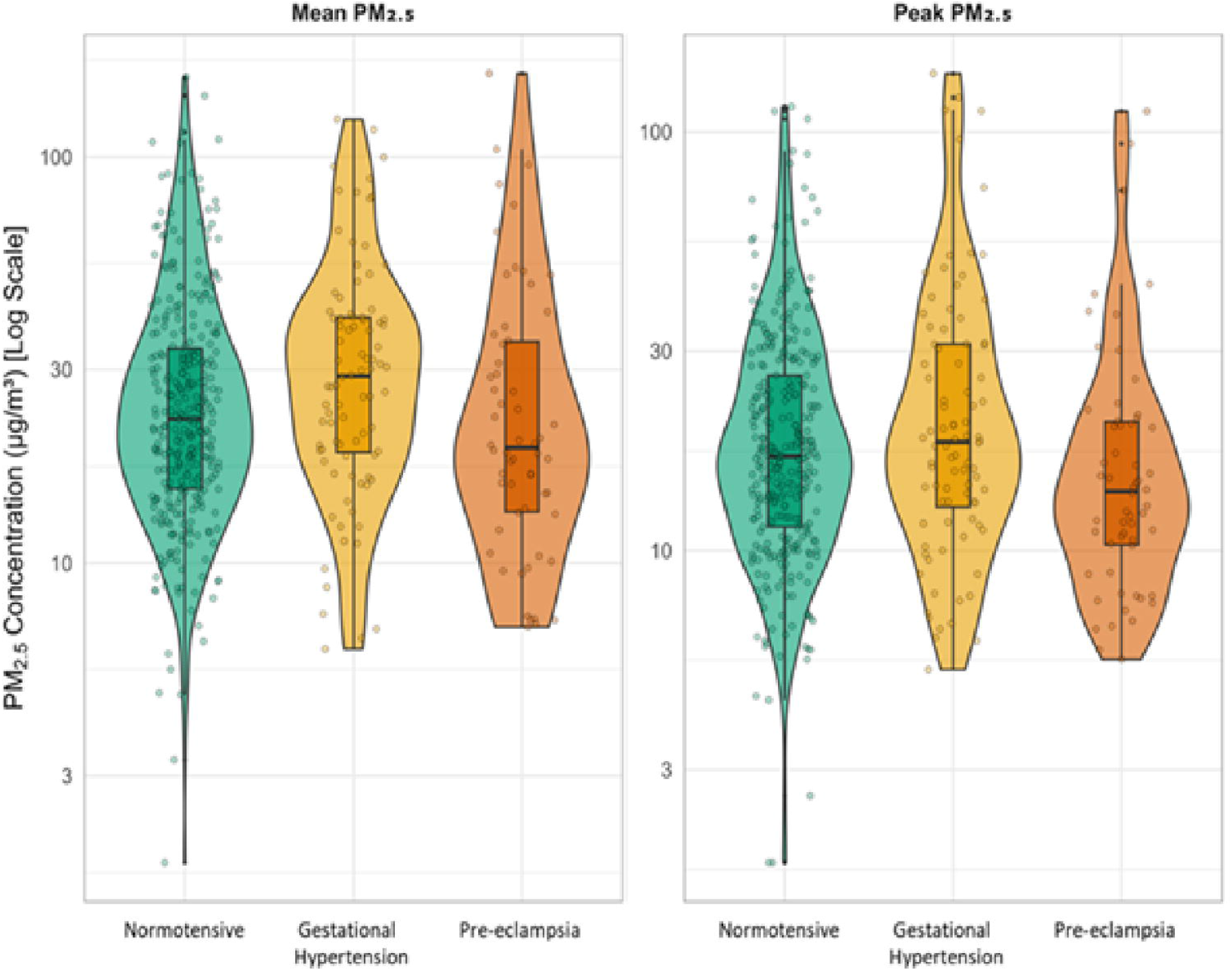

For gestational age at delivery category (figure 2), there was no significant variation detected in mean (Kruskal-Wallis p = 0·902) or peak (Kruskal-Wallis p = 0·632). PM_2·5_ concentrations. Neither mean nor peak PM_2·5_exposure were associated with differences in gestational age at delivery overall, by setting, or by season (all linear regression and GAM p-values > 0·05) (supplementary figures 2 a-c).

**Fig. 2.**
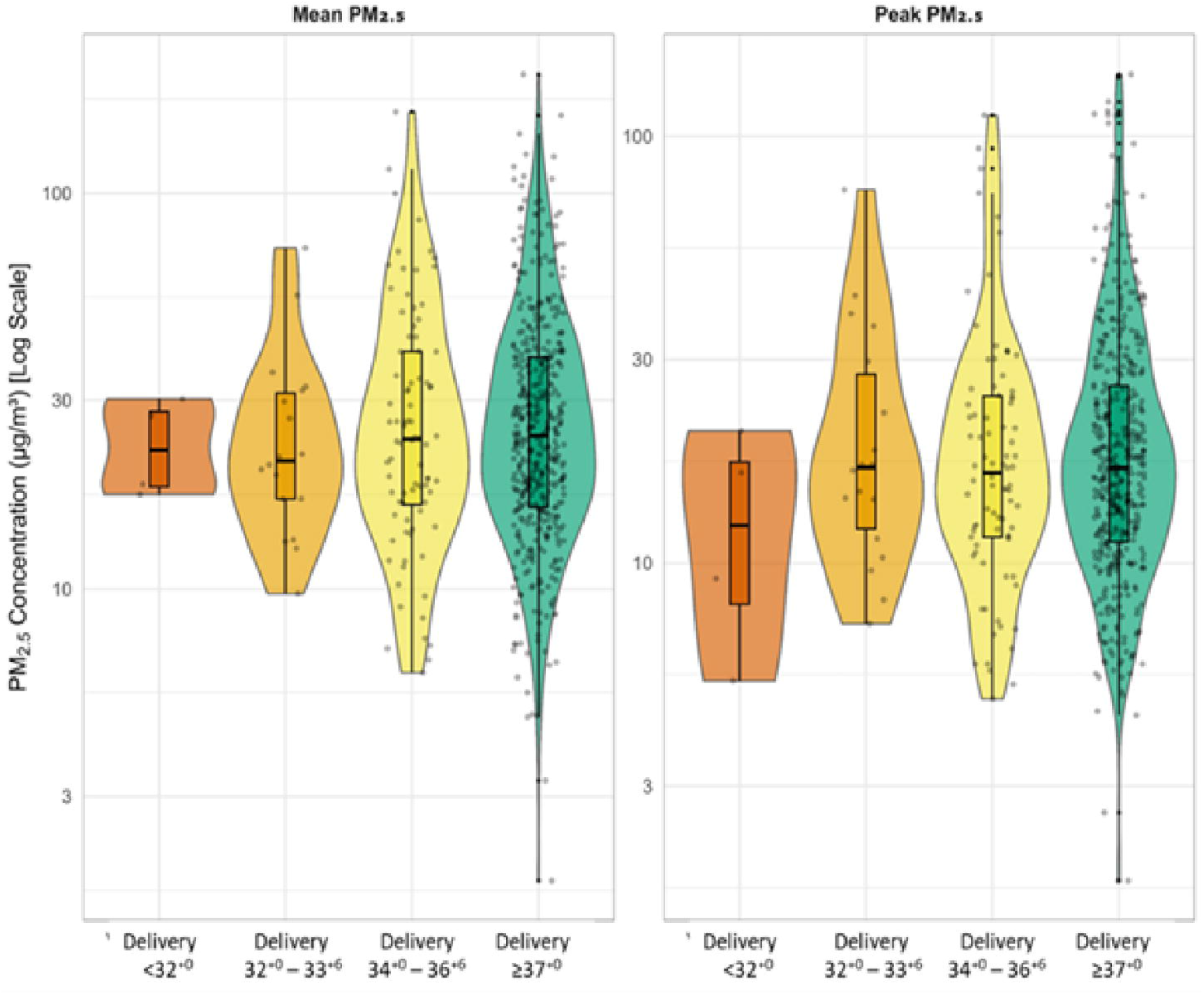

In the analyses of fetal growth velocity (figure 3), mean (Kruskal-Wallis p = 0·032) and peak (Kruskal-Wallis p = 0.047) PM□.□ exposure varied across birthweight categories, with higher mean exposures among SGA compared with LGA infants (Dunn’s p = 0·024). Mean (p = 0·017) and, possibly, peak (p = 0·051) PM□.□ exposures were inversely associated with birthweight centile (supplementary figure 3a). Across rural and urban settings, and during dry seasons, increasing mean and peak PM□.□ concentrations were consistently associated with lower birthweight centiles (supplementary figures 3b and 3c). No differences in PM□.□ concentrations were observed between livebirths and stillbirths (mean: p = 0·441; peak: p = 0·266) (figure 4).

**Fig. 3.**
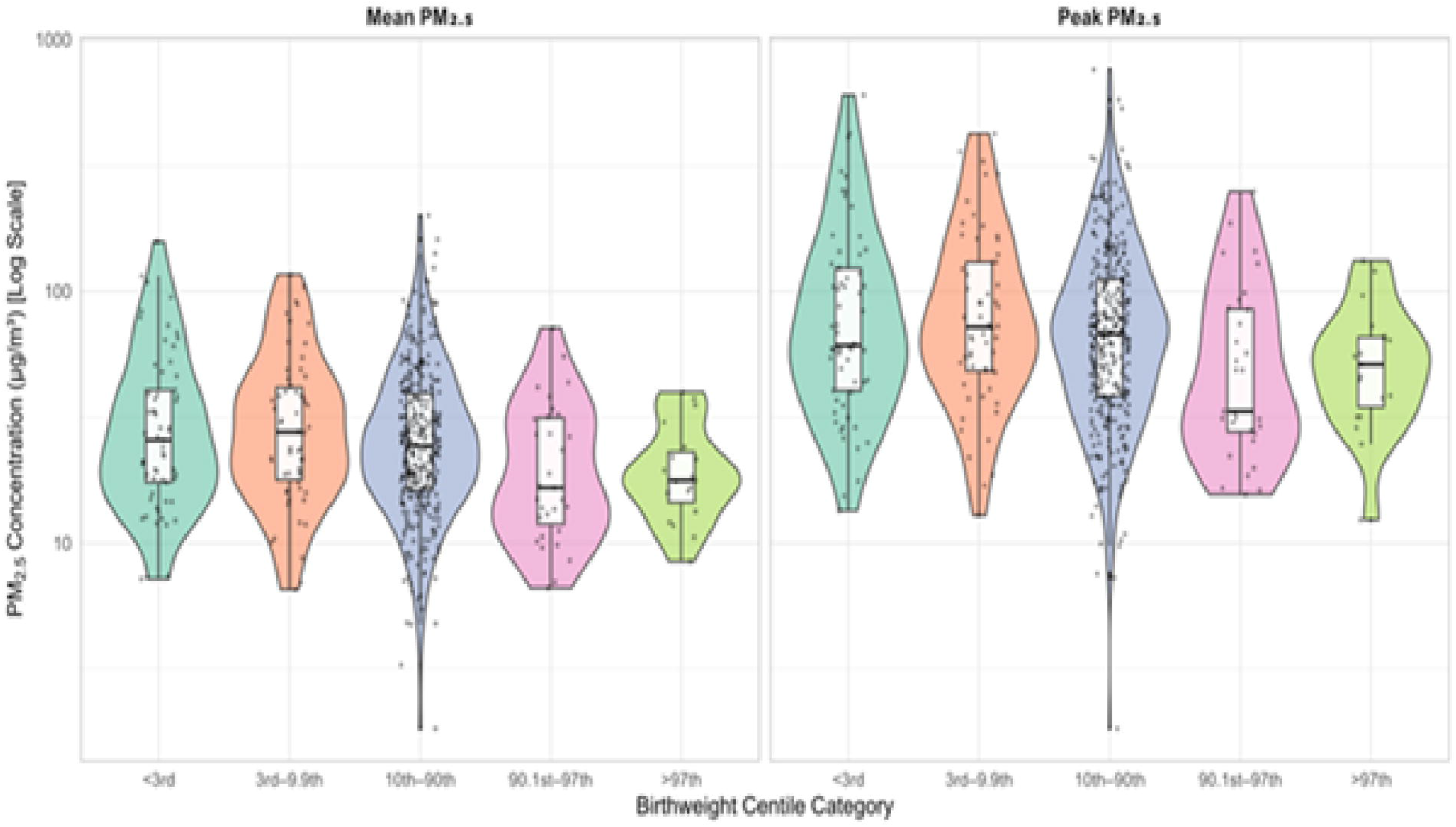

**Fig. 4.**
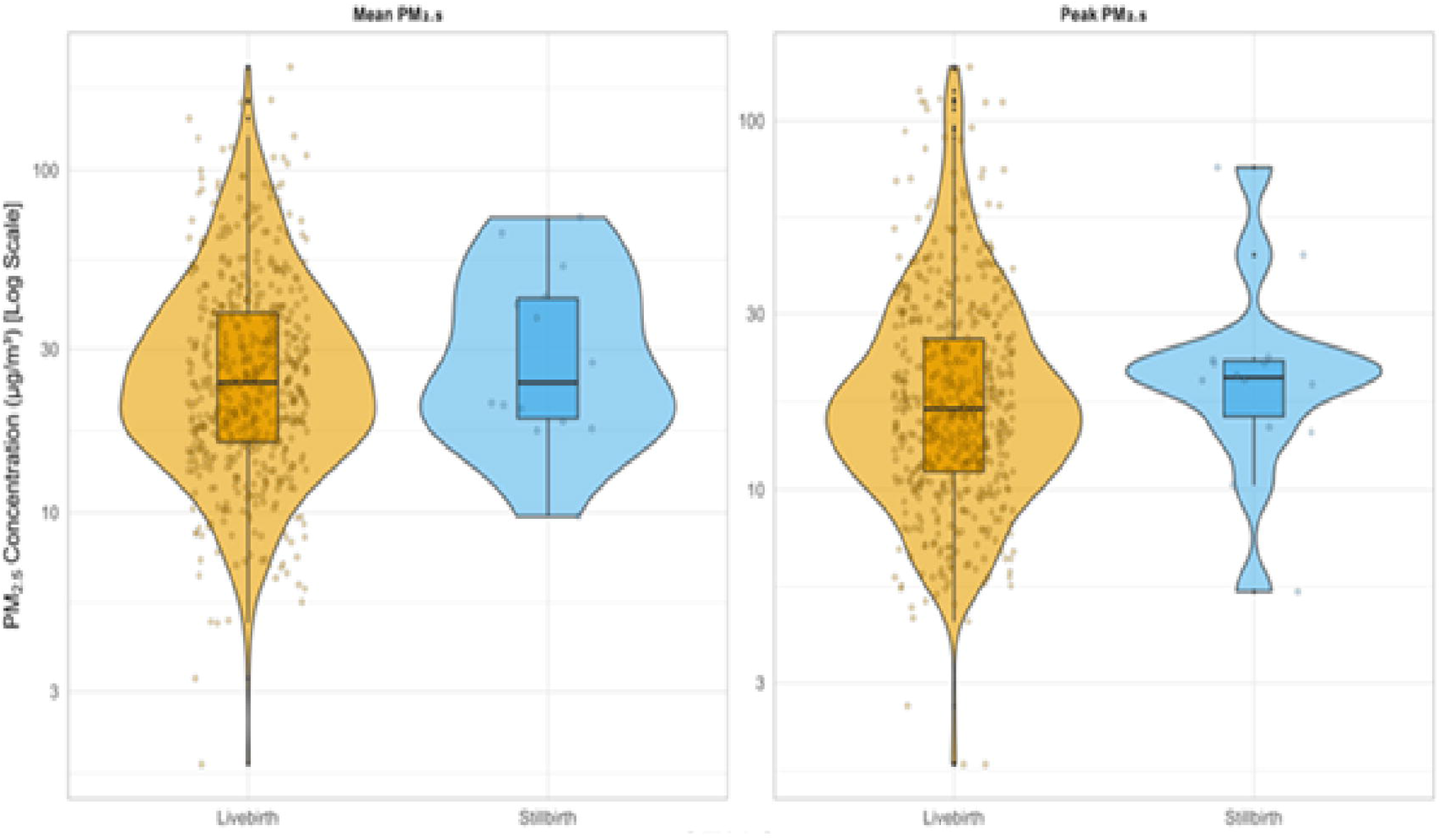

## Discussion

This study provides novel evidence linking directly measured personal exposure to fine particulate air pollution (PM□.□) with placenta-mediated pregnancy complications in three sub-Saharan African countries. Using high-resolution personal monitoring, we found that higher PM□.□ exposure was associated with indicators of impaired fetal growth, particularly during the wet season. These associations were not observed for preterm birth or stillbirth.

Our findings underscore the significant pollution burden experienced by pregnant women in sub-Saharan Africa, with personal PM□.□ exposures exceeding WHO air quality guidelines by substantial margins across all study sites.(20) The magnitude and distribution of these exposures — including extreme peak values frequently surpassing 1000 µg/m^3^ — highlight the pervasive nature of both chronic and acute pollution episodes during pregnancy. These levels are likely to reflect daily mobility through polluted microenvironments such as roadsides, markets, and communal cooking areas, as well as seasonal factors that modulate pollution intensity.

The PM□.□ exposure profiles observed in our cohort suggest that short-lived but intense pollution events contribute significantly to total exposure burden, albeit that these exposure profiles were obtained up to a year after the index pregnancy. Such events may be particularly consequential during critical windows of fetal development, potentially amplifying risks of impaired growth or dysregulated maternal haemodynamics. These findings reinforce the need to broaden conventional exposure assessments beyond ambient means, and to capture personal, time-resolved data in vulnerable populations navigating diverse and informal settings.(21)

The strongest association in this study was between increased PM_2·5_ exposure and reduced fetal growth velocity. This is consistent with all 4 umbrella reviews which describe an association between fine particulate matter and either low birthweight(5) or being born small-for-gestational age.(4, 6, 7) Given the shared pathogenesis of gestational hypertension and pre-eclampsia,(8) we are unsure through what biological mechanisms increased exposure to PM_2·5_ might be associated with gestational hypertension, but not pre-eclampsia. This possible finding requires confirmation in other cohorts as it directly conflicts with recent data from Rome and the umbrella reviews, and does not align with the concept that pre-eclampsia is associated with more innate inflammation than is gestational hypertension.(3, 4, 6-8) We did observe that increasing peak exposures were associated with higher mean arterial pressure, especially in the wet season, which is consistent with data from New York State.(22)

There is moderately-strong evidence of a link between air pollution and preterm birth, that was not replicated in this study.(1, 5, 6)

Limitations include the impact of the Covid-19 pandemic causing more than 12-month delays to both completion of the device sling co-design and the field campaigns, precluding the planned contemporaneous personal air quality measurements during index pregnancies. Therefore, we have retrofitted subsequent However, these are the first data to directly link personal maternal PM_2·5_ exposures with pregnancy outcomes. In addition, the recruited women remained living in the home in which they had dwelt during the index pregnancy and elements of their poverty probability index had not changed. Second, we were unable to assess any i societal disruption-related impacts on PM_2·5_ levels caused by the Covid-19 pandemic on the PM_2·5_ exposure-to-outcome relationships. Third, we could not observe the impact of air pollution at different phases of conception and by trimester.(5-7) Fourth, was sample size, especially for uncommon events such as stillbirth for which there is evidence of an association.(5, 6) Visually, our data suggest a possible association between peak exposures and stillbirth (figure 4). Therefore, for these reasons, these data must be considered to be exploratory.

In summary, using directly recorded personal exposure data, we have determined that increased mean and peak exposure to PM_2·5_ is associated with decreased fetal growth velocity. As we have geolocated all homes in the PRECISE cohort, and have detailed information about the construction, lighting, and heating of their homes, cooking arrangements, employment, distance from roads and industry, and ambient exposures, we are in the process of modelling personal air quality and temperature exposomes for all PRECISE participants during their index pregnancy. Future studies need to be linked with accurate measurement of personal exposure to temperature (heat and cold), seasonality, the built environment, vegetation coverage, water and sanitation, and road networks, and investigate and identify biomarkers of risk.(23-25)

## Author Contributions

PvD, BB, CT, LAM, PTM, and LM designed the concept for this air quality study, embedded within the wider PRECISE and PRECISE-DYAD studies. JN, AK, and HJ, supported LM to train and guide the field teams. LM and PvD co-wrote the first draft of the manuscript, and all authors contributed to the revisions and approved the submitted version. M-LV, MM, and AS managed the central data set. LM was responsible for the statistical analyses with support from BB, JNB, AJW, AS, and PvD. All authors had full access to all data in the study and had final responsibility for the decision to submit for publication. LM, PTM, BB, and M-LV and SJEB accessed and verified the data.

## Acknowledgements

We would like to thank all the study participants and their families. We also thank local governments for hosting us and all facility health workers at the study sites and the PRECISE research team for all the diligent work of data and sample collection and laboratory analysis for the study.

## Data Availability Statement

The PRECISE and PRECISE-DYAD data are de-identified participant-level data. As permitted by existing data sharing and collaboration agreements, the data will be available to academically-active entities (eg, universities, non-governmental organisations, and multilaterals), with the PRECISE Principal Investigator (Peter von Dadelszen), or named delegate, as a named co-investigator and within the limits of the informed consent obtained. External and PRECISE network researchers are welcome to request use of data and samples, the scientific potential of which is then reviewed by the Data and Samples Access Committee. We actively promote our preferred model of working, which is to establish collaborations that ensure representative input from country site researchers and local teams (ie, teams who led on recruitment and data/sample collection) in analysis and reporting. This strengthens the relevance of the outputs, as site teams are best placed to interpret the complexities of their context and data. Data requests are directed to the network via an email contact (precise@ kcl.ac.uk) and follow established data request procedures thereafter. There is a schedule of fees for data or sample access. Our data and sample access processes are described in more detail on the study website.(26)

